# Day-to-day dietary variation shapes overnight sleep physiology: a target-trial emulation in 4.8 thousand person-nights

**DOI:** 10.64898/2026.02.17.26346471

**Authors:** Mariya Shkolnik, Gal Sapir, Smadar Shilo, Yeela Talmor-Barkan, Eran Segal, Hagai Rossman

## Abstract

Sleep architecture is essential for metabolic and cardiovascular health, yet the impact of day-to-day dietary variation on objective sleep physiology remains unclear. Using 4.8 thousand person-nights with real-time dietary logs and multi-stage wearable sleep recordings, we examined how prior-day nutrition relates to next-night sleep under free-living conditions. Higher fiber density was associated with increased restorative sleep, including +0.59 pp deep sleep, +0.76 pp REM sleep, −1.35 pp light sleep, and −1.14 bpm lower mean nocturnal heart rate. Greater plant diversity and higher whole-plant food intake were similarly associated with lower nocturnal heart rate (−0.72 to −0.94 bpm). Meal-timing behaviors primarily influenced sleep duration, sleep-onset latency, and autonomic tone: heavier evening meals were associated with +7.7 min longer total sleep time and +0.73 bpm higher nocturnal heart rate. In contrast, short-term variation in macronutrient energy distribution and micronutrient consumption showed no robust associations with sleep outcomes. When analyses were restricted to more extreme dietary contrasts, effect magnitudes increased while remaining directionally consistent. These findings indicate that routine daily dietary choices, particularly plant-forward composition and meal timing, have immediate and measurable effects on objective sleep architecture.

## Introduction

Sleep is a fundamental determinant of cardiometabolic, cognitive, and mental health. High quality sleep, in terms of duration, regularity, and continuity, supports improved insulin sensitivity, lower blood pressure, improved lipid metabolism and appetite regulation; preserves cognitive performance and memory; and maintains emotional balance, mood stability and stress resilience. Conversely, chronic sleep deficiency or poor sleep quality is linked to increased risks of obesity, type 2 diabetes, hypertension, cardiovascular disease, cognitive decline, depression, and anxiety ^1^. Despite this central role in human physiology, the lifestyle factors that shape nightly sleep physiology remain insufficiently characterized. Diet is increasingly recognized as a potential modulator of sleep through intertwined metabolic, hormonal, and circadian pathways, but existing evidence remains fragmented and inconsistent.

Across observational cohorts and small feeding trials, higher diet quality, particularly patterns rich in minimally processed plant foods and fiber, and dairy, has been associated with better sleep duration, efficiency, and continuity, while saturated fat, sugar, and ultra-processed foods correlate with more fragmented sleep ^2–15^. Evidence for meal timing and chrononutrition is mixed: delayed evening meals and longer eating windows have been linked to both improved and impaired sleep outcomes in different populations ^16–27^. Findings on the influence of micronutrient consumption are similarly heterogeneous, with suggestive but inconsistent benefits reported for magnesium ^28–31^, vitamin D ^32,33^, vitamin E ^34^, zinc ^14,35^, and folate ^36^, and limited or conflicting data for B-vitamins ^37–39^ and calcium ^40^.

A major limitation of the existing knowledge is that most studies lack objective sleep staging, high-resolution dietary data, and methodological tools needed to separate causal effects from confounding. Contemporary epidemiologic guidelines emphasize that large observational datasets should emulate key components of a “target trial” ^41^, including explicit confounder control and rigorous causal-inference estimation methods such as inverse-probability weighting and propensity-score modeling ^41–43^. Nutritional epidemiology frameworks highlight the need for transparent exposure definitions, comprehensive confounder adjustment, and robust sensitivity analyses, requirements rarely met in prior nutrition–sleep research ^44–46^.

To address these gaps, we leveraged the Human Phenotype Project (HPP) ^47^, a large-scale longitudinal digital health cohort integrating granular, time-stamped dietary logs with clinically validated home sleep studies. We developed a reproducible causal-inference framework incorporating machine-learning propensity estimation, overlap trimming, inverse-probability weighting, and bootstrap-based uncertainty quantification, following contemporary methodological recommendations ^41,48–54,55^. This framework approximates key features of a target trial in observational data and enables the estimation of associations that align with causal effects of day-to-day nutritional behavior on next-night sleep, provided standard assumptions hold ^41,49^.

Using more than 4,700 person-nights with synchronized dietary and physiological measurements, we systematically evaluated how prior-day variation in nutritional intake and timing influences next-night sleep duration, continuity, staging, and physiology. By combining high-resolution behavioral and physiological data with modern causal-inference methodology, this study provides one of the most comprehensive examinations to date of how acute, prior-day changes in nutritional behavior shape objective sleep architecture in free-living humans.

**Figure 1.**
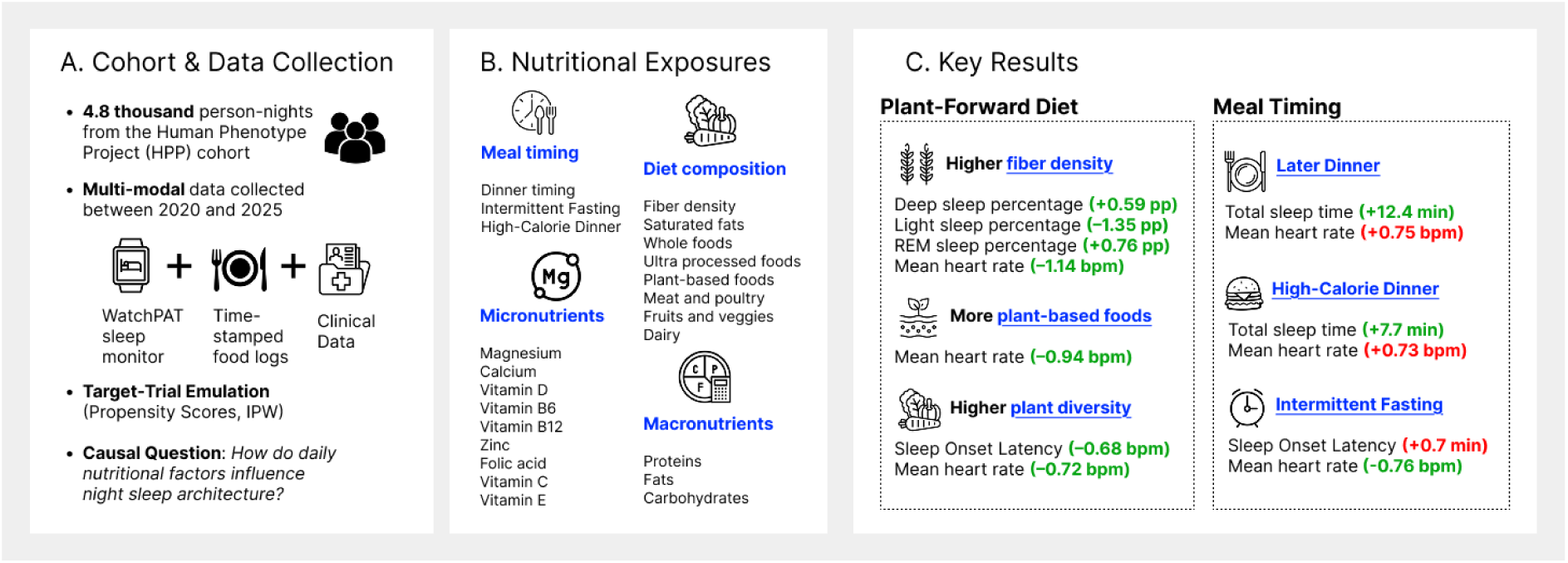
Overview of study design and key findings. The figure provides an overview of the study design, nutritional exposures analyzed, and the two primary “levers” of dietary sleep regulation identified in the Human Phenotype Project (HPP) cohort.

## Results

### Baseline characteristics

We analyzed 4,793 person-nights with concurrent dietary logs and WatchPAT-measured sleep recordings from 3,598 adults enrolled in the HPP, a deeply phenotyped longitudinal cohort in Israel ^47^. The cohort consisted of 1,829 (50.8%) male and 1,769 (49.2%) female participants and included sleep and nutrition data collected between February 2020 and April 2025. Days with implausible total energy intake (<500 or >5,000 kcal), or total sleep duration outside of 300 to 600 minutes were excluded.

Participants were predominantly healthy, with a mean age of 52.7 ± 7.9 years and a mean body mass index (BMI) of 26.0 ± 4.1 kg/m². For more details, see Extended Data Fig. 1 and Extended Data Table 1.

### Data Collection

Sleep was measured under free-living conditions using the WatchPAT 300 device (ZOLL Itamar). Standard indices, including total sleep time, sleep efficiency, arousal burden, and nocturnal heart rate, were derived using manufacturer algorithms ^56^. Additional technical details and validation references are provided in Methods. Daily dietary intake was logged in real time through a custom mobile application, enabling precise capture of meal timing, food type, and portion size ^57^. A full description of dietary logging and nutrient processing is available in the Methods. Daily nutritional exposures were defined as binary high-vs-low contrasts based on population medians, chosen to reflect realistic variation in free-living dietary behavior and to preserve overlap across confounder distributions.

Exposures spanned four domains: diet quality, macronutrient distribution, micronutrient intake, and meal timing. An extended description of all exposure definitions and distributions are provided in the Extended Data Table 2 and Extended Data Table 3 in Appendix.

### Global landscape of nutrition and sleep associations

The analysis included 4,793 logged days from 3,598 participants. On average, participants slept 6.3 hours per night, with sleep composition consisting of 17.6% deep sleep, 24.0% REM sleep, and 58.5% light sleep. Mean sleep efficiency was 88.8%, and average sleep latency was 15 minutes. Mean daily energy intake was 1,815 kcal, with an average macronutrient intake of 80 g protein, 77 g fat, and 200 g carbohydrates, corresponding to 17.6%, 38.4%, and 44% of total energy, respectively.

We evaluated 25 prespecified dietary exposures spanning diet composition, macronutrient distribution, micronutrients, and meal timing for associations with next-night sleep physiology. After correction for multiple hypothesis testing, six exposures showed significant associations with at least one sleep outcome (Fig. 2). The six exposures that remained significant were predominantly plant-forward dietary features (fiber density, plant diversity, whole-plant food intake) and meal timing behaviors.

**Figure 2.**
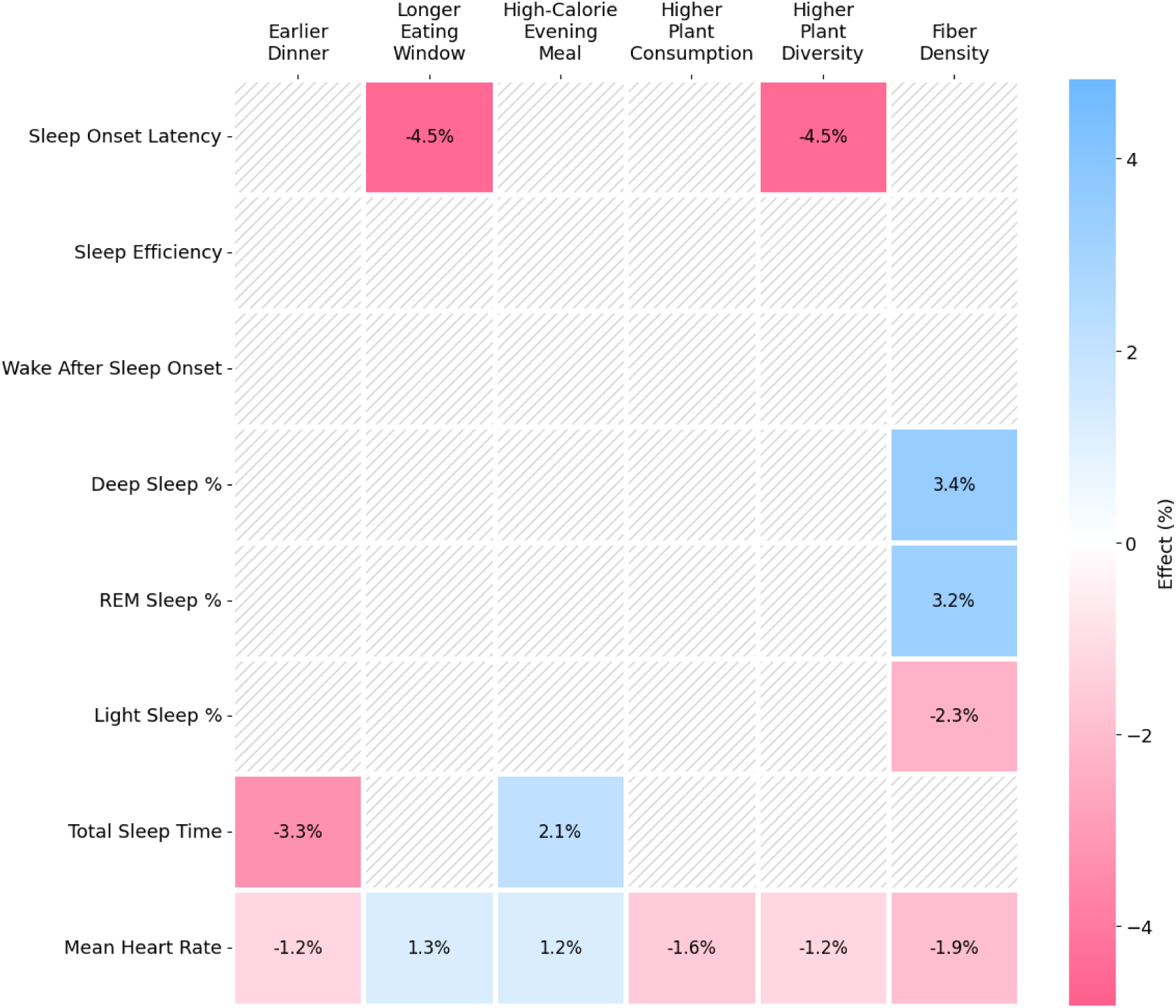
Relative effects of nutritional exposures that significantly influence next-night sleep architecture. Daily nutritional exposures for which the inverse-probability–weighted average treatment effect (ATE) was statistically significant after bootstrap-based uncertainty estimation and false discovery rate (FDR) correction across prespecified sleep outcomes. Effect sizes represent the percent difference in each sleep outcome between higher- and lower-intake days (Hajek-normalized IPW). Blue indicates higher values in the higher-intake group, red indicates lower values, and hatched cells mark outcome-exposure pairs that did not reach significance. Meal timing effects were observed for sleep duration, sleep onset latency, and nocturnal heart rate, whereas diet composition effects were concentrated on sleep-stage distribution and nocturnal autonomic physiology.

### Meal timing

Meal-timing behaviors primarily influenced sleep duration, sleep-onset latency, and autonomic tone, while leaving sleep-stage composition largely unaffected. Higher in calories evening meals were associated with longer sleep duration and elevated nocturnal heart rate, whereas earlier dinner timing was linked to shorter sleep duration and lower heart rate. Longer eating windows were associated with prolonged sleep-onset latency and higher nocturnal heart rate. These findings suggest that the temporal distribution of food intake acts as a key modulator of sleep quantity and cardiovascular regulation. The relative effects of these timing behaviors are illustrated in Figure 3.

**Figure 3.**
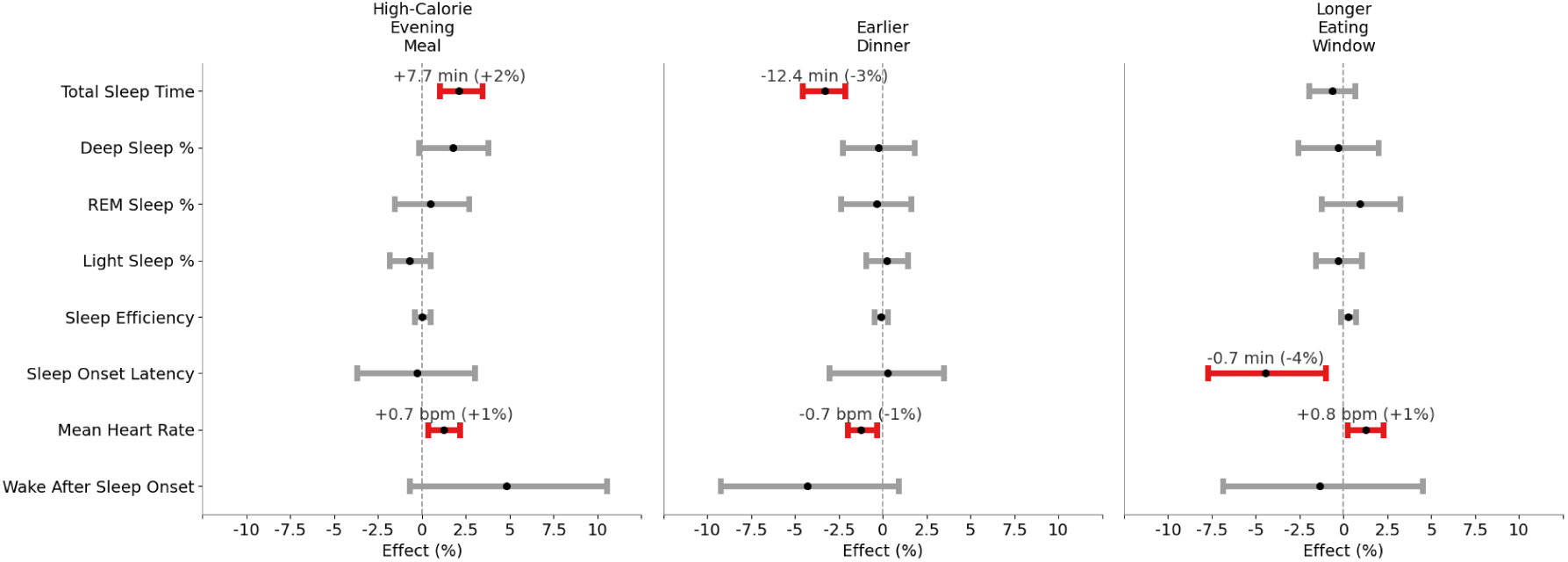
Relative effects of daily meal-timing behaviors on next-night sleep physiology. Shown are inverse-probability–weighted average treatment effects (ATEs), expressed as percent differences between higher- and lower-exposure days, for three meal-timing behaviors: high-calorie evening meals, earlier dinner timing, and longer daily eating windows. Effect estimates were obtained using Hajek-normalized inverse probability weighting, with uncertainty assessed by nonparametric bootstrapping and significance determined after false discovery rate (FDR) correction across prespecified sleep outcomes. Red markers indicate statistically significant effects after FDR correction; grey markers denote non-significant exposure–outcome pairs. Outcomes include sleep duration, sleep-stage composition, sleep continuity, and mean heart rate during sleep.

A longer daily eating window was associated with a higher mean sleeping heart rate on the target night (+0.76 bpm, 95% CI [+0.13, +1.33]; p = 0.048) and shorter sleep onset latency (−0.67 min, 95% CI [−1.17, −0.15]; p = 0.048). Earlier dinner timing was associated with shorter total sleep time on the target night (−12.39 min, 95% CI [−17.13, −7.98]; p < 0.01) and lower mean sleeping heart rate (−0.74 bpm, 95% CI [−1.23, −0.21]; p = 0.04). A heavier evening meal was associated with longer total sleep time (+7.73 min, 95% CI [+3.71, +12.37]; p < 0.01) and a modest increase in mean sleeping heart rate (+0.73 bpm, 95% CI [+0.19, +1.26]; p = 0.032).

### Diet Composition

Dietary composition effects were concentrated on sleep-stage distribution and nocturnal heart rate rather than sleep duration. Higher fiber density was associated with a more restorative sleep architecture, characterized by increased proportions of deep and REM sleep and a reduction in light sleep. Additionally, plant-forward patterns, including greater plant diversity and higher whole-plant food intake, were associated with improved nocturnal autonomic down-regulation. These associations between diet quality and objective sleep architecture are summarized in Figure 4.

**Figure 4.**
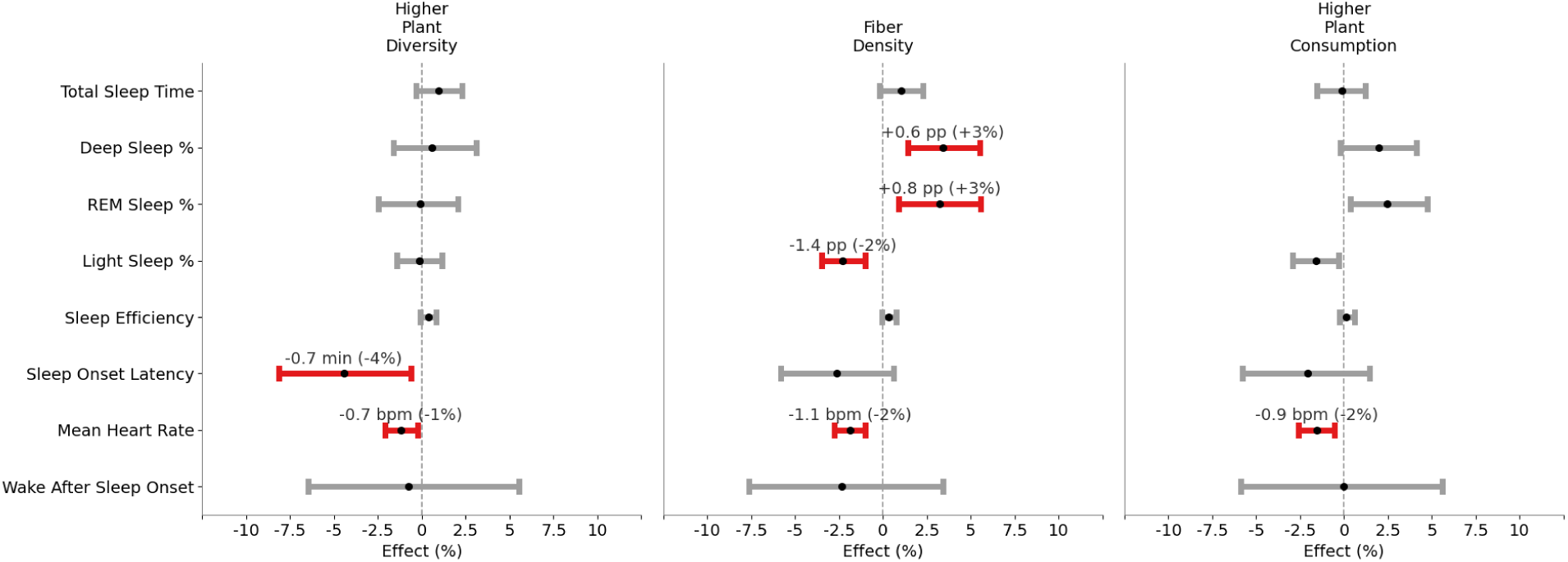
Relative effects of diet-quality components on next-night sleep physiology. Shown are inverse-probability–weighted average treatment effects (ATEs), expressed as percent differences between higher- and lower-exposure days, for diet-composition features that remained significant after false discovery rate (FDR) correction. Effect estimates were derived using Hajek-normalized inverse probability weighting with bootstrap-based uncertainty estimation. Grey markers denote exposure–outcome pairs that did not reach statistical significance after correction. Outcomes include sleep-stage proportions, sleep continuity metrics, total sleep time, and mean heart rate during sleep.

Higher fiber density was associated with a lower mean sleeping heart rate on the target night (−1.14 bpm, 95% CI [−1.67, −0.60]; p < 0.01). In parallel, fiber density was associated with structured redistribution of sleep stages: deep sleep increased by +0.59 percentage points (95% CI [+0.25, +0.94]; p = 0.008), REM sleep increased by +0.76 percentage points (95% CI [+0.21, +1.29]; p = 0.008), and light sleep decreased by −1.35 percentage points (95% CI [−2.08, −0.59]; p < 0.01). Greater plant diversity was associated with shorter sleep onset latency (−0.68 min, 95% CI [−1.27, −0.09]; p = 0.048) and lower mean sleeping heart rate (−0.72 bpm, 95% CI [−1.27, −0.15]; p = 0.048). A higher whole-plant foods ratio was associated with a lower mean sleeping heart rate on the target night (−0.94 bpm, 95% CI [−1.56, −0.32]; p = 0.016). Nominal associations with reduced light sleep and increased REM sleep did not remain significant after FDR correction (p = 0.064 for both).

### Sensitivity analyses and negative controls

We performed a comprehensive robustness assessment across all 25 exposure models. Covariate balance was evaluated using the absolute standardized mean difference (ASMD), with a prespecified threshold of 0.1 for acceptable mean balance, consistent with established recommendations ^58^. To assess whether observed effects could reflect generalized modeling artifacts rather than nutrition-specific associations, we analyzed three food-insensitive negative-control outcomes: heart-rate signal-quality score, percent supine sleep, and left-side sleep duration. Across 1,000 bootstrap replications per exposure, none of the negative-control outcomes showed statistically significant associations (0% false-positive rate). To further validate the causal-inference framework, we conducted a positive-control analysis focusing on caffeine timing. As expected, shorter intervals between the last caffeinated beverage and bedtime were associated with a significant shift toward lighter sleep architecture, including reduced light sleep percentage and increased sleep efficiency. This consistency with established physiological effects of caffeine provides additional support for the internal validity and sensitivity of our modeling pipeline. Detailed diagnostics and positive-control analysis are reported in the Methods. Together, adequate covariate balance, null findings in negative controls, and an expected effect in a positive control support the interpretation that the reported associations reflect reproducible structure in the data rather than modeling artifacts.

## Discussion

This study provides a comprehensive evaluation of how acute, day-to-day nutritional choices influence objective sleep architecture and nocturnal autonomic regulation in a large, free-living cohort. By integrating granular dietary logs with multi-stage wearable sleep recordings, we identify distinct roles for diet composition and meal timing as primary modulators of next-night sleep physiology.

Our primary findings demonstrate that meal-timing behaviors significantly influence sleep quantity and autonomic tone. Specifically, higher calorie evening meals were associated with increased total sleep duration but elevated nocturnal heart rates, whereas earlier dinner timing correlated with shorter sleep and improved cardiovascular regulation. Longer daily eating windows were linked to reduced sleep-onset latency and elevated nocturnal heart rate. These findings parallel ongoing controversies in the chrononutrition field, where effects appear to depend strongly on context, health status, and prior metabolic load ^18,21,25,27^. Although chrononutrition research emphasizes earlier meal timing for metabolic health ^20,21,26^, several human studies suggest that moderately shorter intervals between the last meal and sleep, or carbohydrate-rich evening meals, can acutely prolong total sleep duration or reduce sleep onset latency ^16,17,19,22–24,26,27^.

In contrast to meal timing, diet composition effects were concentrated on sleep-stage distribution and nocturnal heart rate. Higher fiber density was associated with increased deep and REM sleep proportions and reduced light sleep, alongside lower nocturnal heart rate, extending controlled-feeding evidence that fiber-rich meals enhance slow-wave physiology ^5–8,12^. Greater plant diversity and higher whole-plant food intake were associated with shorter sleep-onset latency and reduced nocturnal heart rate, suggesting improved nocturnal autonomic down-regulation ^2,4,15^. These patterns suggest that plant-forward dietary features may influence sleep quality through pathways distinct from timing, potentially involving gut–brain signaling, inflammatory tone, or metabolic stability. Together, these findings suggest that temporal aspects of food intake modulate sleep quantity and autonomic tone, rather than altering sleep-stage allocation.

While most individual micronutrients did not remain statistically significant after false discovery rate correction across the full exposure set, several plant-associated vitamins and minerals, including magnesium and folic acid, exhibited directionally consistent associations with sleep-stage distribution and nocturnal heart rate, with confidence intervals that did not cross zero for specific outcomes (Supplementary Results Fig. S1). These effects were modest in magnitude and did not generalize uniformly across all sleep metrics, precluding strong inference about isolated nutrient effects. These patterns are more plausibly interpreted within a food-matrix framework, in which micronutrients act as correlated markers of broader dietary contexts rather than as independent drivers of physiology. In this view, plant-derived nutrients may reflect composite exposures that include fiber content, polyphenols, fermentable substrates, and meal structure, all of which may jointly influence metabolic and autonomic pathways relevant to sleep.

A central contribution of this work lies in its integration of objective sleep metrics with a high-dimensional causal inference framework. First, it leverages one of the largest datasets to date linking detailed, timestamped dietary intake with objective, multi-stage sleep physiology measured under free-living conditions. Second, the use of a modern causal-inference framework, including machine-learning propensity estimation, overlap trimming, and bootstrap-based uncertainty, enables causal interpretation while adhering to current epidemiologic guidelines for target-trial emulation. Third, the approach examines realistic behavioral contrasts, using median-based exposure definitions that reflect common habits rather than extreme diets. Finally, the inclusion of negative-control outcomes further supports internal validity by showing no spurious associations for features unrelated to diet.

Several limitations warrant consideration. Dietary data are self-reported and subject to under-logging or recall bias, although time-stamping reduces misclassification. Day-level exposures may obscure dose-response relationships or timing-specific nuances (e.g., morning vs. evening intake within a macronutrient class). The median-split exposure design simplifies interpretation but may mask non-linearities and shifts attributable to food sources, especially in composite categories like whole foods. Although extensive confounder adjustment was performed, residual confounding from unmeasured factors on a certain day, stress, and irregular routines, remains possible. Finally, our findings describe acute, next-night effects and do not address chronic adaptation or long-term dietary patterns.

In conclusion, in one of the largest real-world analyses integrating high-resolution dietary logs with objective sleep staging, we demonstrate that everyday nutritional behaviors significantly affect sleep physiology. Fiber intake, plant-forward dietary patterns, were consistently linked to more restorative sleep architecture. Meal timing emerged as an independent modulator of sleep quantity and continuity. The study shows that even modest day-to-day changes in dietary patterns, like increasing fiber or shifting meal timing, can have measurable effects on sleep outcomes. This suggests that small and practical dietary tweaks might be leveraged to improve sleep quality. The heterogeneity observed across exposures suggests that individuals may differ meaningfully in their day-level sensitivity to specific dietary behaviors. Future work should validate and quantify these heterogeneous treatment effects using richer stratification (e.g., metabolic phenotype, chronotype, habitual diet), which may clarify why certain individuals exhibit stronger sleep responses than others.

## Methods

### Study population

Data was derived from the HPP, a longitudinal health cohort launched at the Weizmann Institute of Science. The cohort represents one of the largest ongoing population studies in Israel, composed primarily of community-dwelling adults of diverse but predominantly European (Ashkenazi) ancestry, residing in a geographically and environmentally homogeneous region. Participants were generally healthy at enrollment, with severe chronic illnesses excluded at recruitment. Repeat visits occur biennially over a planned 25-year follow-up. For full study design, see ^47^.

### Sleep assessment

Sleep was assessed using the WatchPAT 300 (ZOLL Itamar Medical), an FDA-cleared and clinically validated home sleep test. The device measures peripheral arterial tone, heart rate, oxygen saturation, snoring, and classifies sleep into REM, deep, and light stages via automated proprietary algorithms. For a full description of the device pipeline, validation metrics, and sleep event processing, see ^56^.

Although 7-9 h of sleep is recommended for optimal adult health ^59^, we restricted our sample to nights with 5-10 h of sleep, as an adequate range for single-night sleep to capture typical variability while excluding extreme durations ^60^. For full details of the data selection, see Extended Data Fig. 1. To capture nutrition-sensitive physiology, we focused on a set of eight key outcomes capturing core domains of sleep physiology and autonomic regulation. These included total sleep time as a direct measure of sleep duration, which prior literature shows is sensitive to dietary composition and meal timing ^61^, measures of sleep stage architecture, specifically percent deep, REM, and light sleep, which can vary with macronutrient intake and metabolic processes linked to diet ^61^; and sleep continuity metrics such as sleep efficiency, sleep onset latency, and wake time after sleep onset, which have been associated with pre-sleep meal characteristics and macronutrient balance in controlled studies ^62^. We also incorporated mean sleeping heart rate to reflect overall autonomic state during sleep, given evidence that meal timing and chrononutrition modulate nocturnal cardiovascular regulation ^63^. Three positional and signal-quality metrics served as negative-control outcomes.

### Dietary assessment and exposure definitions

Dietary intake was logged in real time through a custom HPP mobile application, in which participants recorded meal timing, ingredients, and portion size. Logs were mapped to a comprehensive nutrient database derived from the Israeli Ministry of Health dataset and extended with curated international sources ^57^. Daily totals were computed for energy, macronutrients, micronutrients, food-quality indices, and meal-timing variables. Automated QC removed implausible entries and dense logging artifacts. Only days with >500 and <5,000 kcal kcal logged and a corresponding valid sleep recording were included. For full details of the data selection, see Extended Data Fig. 1. For each exposure, the high/low threshold was set using the median (or sex-specific median for micronutrients) of the baseline distribution, and then applied to the corresponding target-day intake to define ‘high’ vs. ‘low’ exposure. This approach created balanced groups, ensured propensity-score overlap, and reflected realistic behavioral contrasts rather than extreme diets. Full definitions and distributions of all exposure variables are provided in the Extended Data Table 2 and Extended Data Table 3.

### Covariates

To estimate the acute, quasi-causal effects of daily nutrition, we implemented a longitudinal lagged-variable design. This architecture utilizes a clear temporal sequence to minimize confounding from stable individual traits and prior behavioral states:

- Baseline State (Day N): we utilized dietary intake (macronutrients, micronutrients, and timing) and objective sleep architecture from the preceding day as primary confounders.
- Exposure (Day N+1): The nutritional “treatment” was defined based on dietary choices made during the next day.
- Outcome (Night N+1): Sleep architecture and autonomic physiology were measured on the night immediately following the exposure.

By adjusting for Day N sleep and nutrition, we ensure that the estimated effects of Day N+1 dietary choices are controlled for the participant’s immediate baseline physiological state and habitual intake. Additional time-varying and fixed covariates included demographics (age, sex, BMI), lifestyle behaviors (physical activity, smoking, alcohol, caffeine intake/timing), medical history, and daily mood/stress indicators. Analyses were strictly restricted to participant-days with complete data for both the baseline (Day N) and target (Day N+1) periods to ensure proper temporal ordering. To adjust for confounding, we trained exposure-specific propensity score (PS) models.

### Causal Inference Framework

To estimate quasi-causal effects of daily nutritional exposures on next-night sleep, we implemented a modern epidemiologic framework aligned with target-trial emulation and current recommendations for propensity-score–based weighting ^41,48–54^ ^55^.

For each exposure, we fit a separate propensity-score (PS) model to estimate the probability of receiving the high vs. low exposure. PS were estimated using a CatBoost gradient-boosted tree classifier trained on a comprehensive set of baseline and time-varying covariates. To avoid information leakage across nights from the same participant, we performed a participant-level split: unique individuals were randomly partitioned into training (70%) and test (30%) sets, and all nights from a given individual were assigned to the same subset. Class imbalance between treated and control nights was handled via inverse-frequency class weights when fitting CatBoost. Predicted probabilities from the fitted CatBoost model were then calibrated using Platt scaling ^52^. For each exposure, a single calibrated PS model was estimated and then used for all subsequent trimming and weighting configurations in sensitivity analyses.

Analyses were restricted to regions of common support in the PS distribution. For each exposure, we computed the minimum and maximum PS within the treated and control groups and retained observations whose PS lay in the intersection of these ranges. Within this overlap region, we additionally applied one-sided quantile trimming to down-weight extreme PS values: treated observations with very low PS (below the q-quantile among treated) and control observations with very high PS (above the (1-q)-quantile among controls) were excluded. Trimming thresholds q were selected separately for each exposure from a prespecified range in order to optimize covariate balance and preserve adequate sample size ^51,64^.

We then constructed inverse-probability weights using a Hájek-type estimator. For each bootstrap replicate (see below), we computed weights

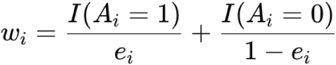

stabilized by the marginal treatment prevalence and clipped at upper and lower percentiles to reduce the influence of extreme values. Within each arm, weights were Hájek-normalized, which yields stable finite-sample estimates of the arm-specific means.

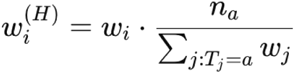

Average treatment effects (ATEs) were computed as the difference between weighted mean outcomes in the treated and control groups and reported both in absolute units and as percent differences relative to the control mean ^54^.

Uncertainty was quantified using a nonparametric bootstrap applied to the full IPW pipeline. For each exposure, we drew 1,000 bootstrap samples with replacement, recomputed weights and ATEs in each replicate, and derived percentile-based 95% confidence intervals from the empirical bootstrap distribution. Two-sided p-values were computed. All analyses were conducted strictly within the overlap and trimmed region for each exposure, avoiding extrapolation to unsupported parts of the covariate and propensity-score space.

### Robustness and negative-control analyses

For balance diagnostics, we focused on a prespecified subset of primary confounders, including (i) structural variables (age, sex, and body mass index), (ii) baseline sleep characteristics, and (iii) the top contributors to the propensity score (PS) model, defined as 15 covariates with the highest contribution to PS estimation. While all available confounders were included in PS model training, balance assessment was restricted to these key variables to ensure interpretability and to prioritize covariates most influential for treatment assignment and outcome prediction. For each exposure, covariate balance was assessed using the absolute standardized mean difference (ASMD). Acceptable balance was defined a priori using an ASMD threshold of ≤0.10 for all covariates, consistent with established recommendations ^58^. For key structural confounders (age, sex, and body mass index) and confounders reflecting baseline sleep characteristics, we applied a more stringent prespecified threshold of ≤0.05, evaluated across all bootstrap replications.

We considered exposure-specific results reliable when three conditions were met: (i) ATE estimates were stable across reasonable trimming and weighting configurations, (ii) weighted ASMDs demonstrated good balance for both structural and data-driven confounders, and (iii) negative-control outcomes exhibited no evidence of systematic treatment effects ^55^. As shown in Table 1, all exposure models satisfied these criteria, supporting the robustness of the reported associations.

**Table 1.**
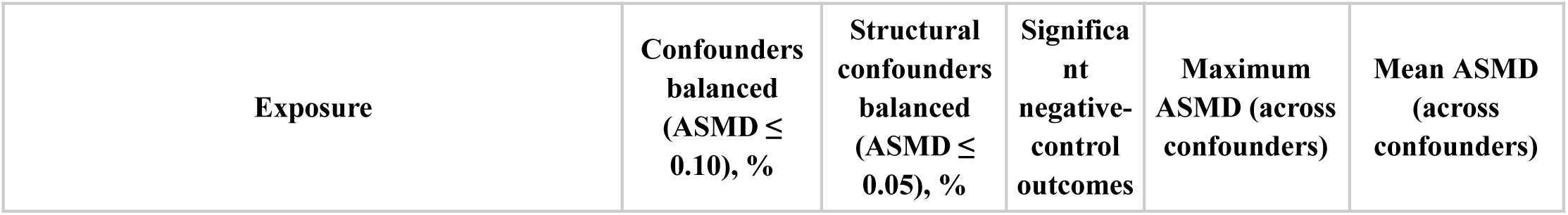
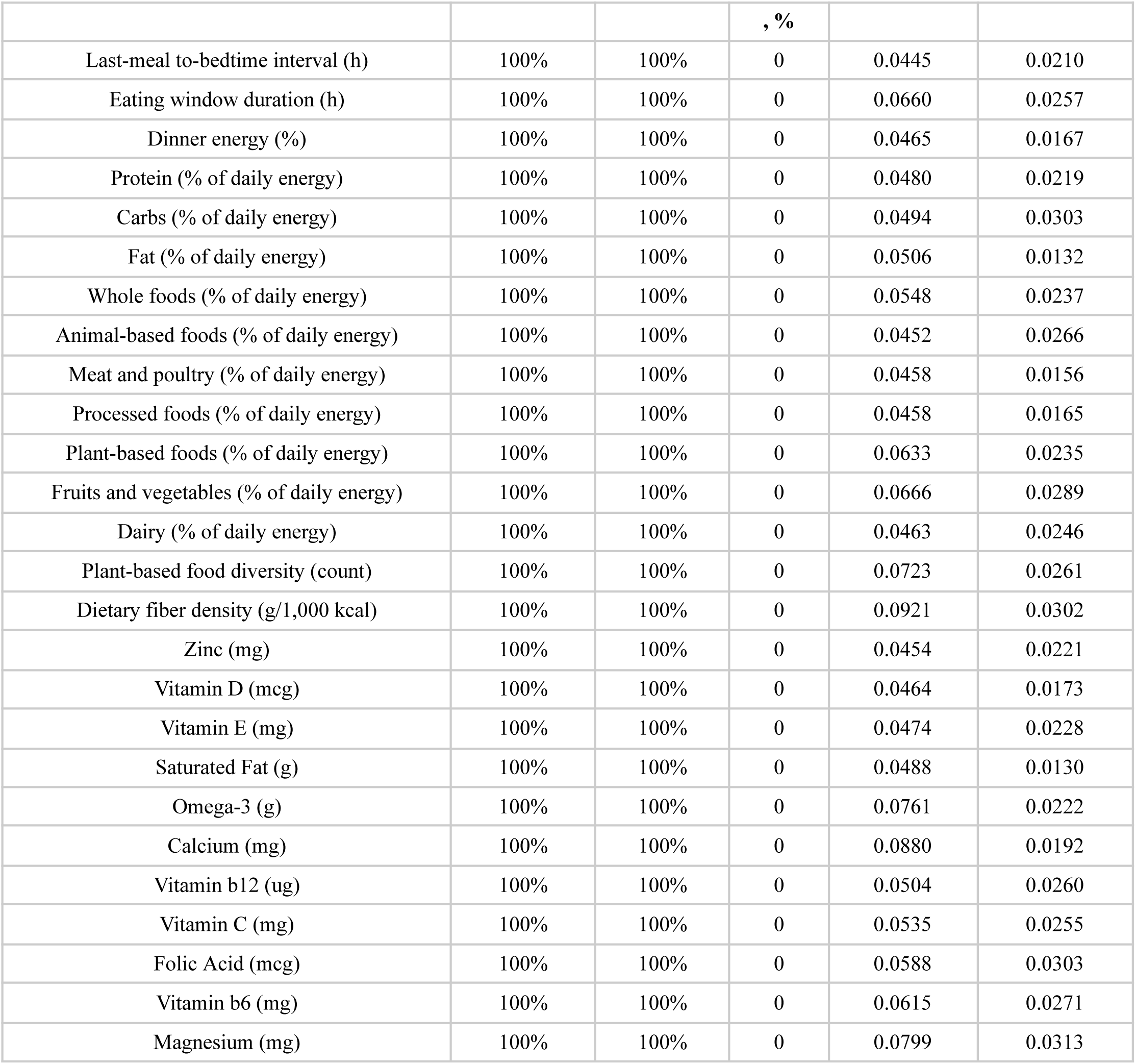
Covariate balance and robustness diagnostics for each exposure model. For each exposure, covariate balance was assessed across 1,000 bootstrap replications using the absolute standardized mean difference (ASMD) for all main confounders (n ≥ 15). Reported values summarize balance diagnostics across bootstrap iterations, including the proportion of confounders meeting prespecified ASMD thresholds, as well as the mean and maximum ASMD across all confounders and bootstrap samples. Negative-control outcomes were evaluated using two-sample t-tests applied to weighted data to assess residual systematic differences between treated and control groups.

| Exposure | Confounders balanced (ASMD $\leq 0.10$), % | Structural confounders balanced (ASMD $\leq 0.05$), % | Significant negative-control outcomes | Maximum ASMD (across confounders) | Mean ASMD (across confounders) |
| --- | --- | --- | --- | --- | --- |

*Table 1. Covariate balance and robustness diagnostics for each exposure model.*
|  |  |  | , % |  |  |
| --- | --- | --- | --- | --- | --- |
| Last-meal to-bedtime interval (h) | 100% | 100% | 0 | 0.0445 | 0.0210 |
| Eating window duration (h) | 100% | 100% | 0 | 0.0660 | 0.0257 |
| Dinner energy (%) | 100% | 100% | 0 | 0.0465 | 0.0167 |
| Protein (% of daily energy) | 100% | 100% | 0 | 0.0480 | 0.0219 |
| Carbs (% of daily energy) | 100% | 100% | 0 | 0.0494 | 0.0303 |
| Fat (% of daily energy) | 100% | 100% | 0 | 0.0506 | 0.0132 |
| Whole foods (% of daily energy) | 100% | 100% | 0 | 0.0548 | 0.0237 |
| Animal-based foods (% of daily energy) | 100% | 100% | 0 | 0.0452 | 0.0266 |
| Meat and poultry (% of daily energy) | 100% | 100% | 0 | 0.0458 | 0.0156 |
| Processed foods (% of daily energy) | 100% | 100% | 0 | 0.0458 | 0.0165 |
| Plant-based foods (% of daily energy) | 100% | 100% | 0 | 0.0633 | 0.0235 |
| Fruits and vegetables (% of daily energy) | 100% | 100% | 0 | 0.0666 | 0.0289 |
| Dairy (% of daily energy) | 100% | 100% | 0 | 0.0463 | 0.0246 |
| Plant-based food diversity (count) | 100% | 100% | 0 | 0.0723 | 0.0261 |
| Dietary fiber density (g/1,000 kcal) | 100% | 100% | 0 | 0.0921 | 0.0302 |
| Zinc (mg) | 100% | 100% | 0 | 0.0454 | 0.0221 |
| Vitamin D (mcg) | 100% | 100% | 0 | 0.0464 | 0.0173 |
| Vitamin E (mg) | 100% | 100% | 0 | 0.0474 | 0.0228 |
| Saturated Fat (g) | 100% | 100% | 0 | 0.0488 | 0.0130 |
| Omega-3 (g) | 100% | 100% | 0 | 0.0761 | 0.0222 |
| Calcium (mg) | 100% | 100% | 0 | 0.0880 | 0.0192 |
| Vitamin b12 (ug) | 100% | 100% | 0 | 0.0504 | 0.0260 |
| Vitamin C (mg) | 100% | 100% | 0 | 0.0535 | 0.0255 |
| Folic Acid (mcg) | 100% | 100% | 0 | 0.0588 | 0.0303 |
| Vitamin b6 (mg) | 100% | 100% | 0 | 0.0615 | 0.0271 |
| Magnesium (mg) | 100% | 100% | 0 | 0.0799 | 0.0313 |

As an additional validation, we conducted a positive-control analysis. Randomized controlled trials have consistently demonstrated that caffeine consumption impairs sleep quality, particularly when consumed closer to bedtime ^65,66,67^. We applied our pipeline to the interval between the last caffeinated beverage and bedtime, comparing 3.6h (±1.5) versus 10.9h (±3.1) intervals between the last caffeinated beverage intake to bedtime intervals. In alignment with established evidence ^65,66,67^, shorter intervals between caffeine consumption and sleep were associated with a significant shift toward lighter sleep architecture. Specifically, we observed a significant increase in light sleep percentage (+2.1 pp; p=0.048) and a decrease in sleep efficiency (−0.8 pp, p=0.048) in a group taking caffeine closer to bedtime, demonstrating the sensitivity of our framework to known lifestyle-sleep disruptions. Collectively, the achievement of adequate covariate balance, the null findings for negative controls, and the concordance of the positive control with known causal effects support the robustness of our reported associations.

### Software and reproducibility

All analyses were conducted in Python, using CatBoost, NumPy, Pandas, and SciPy libraries. Code for the causal-inference pipeline, preprocessing steps, and figure generation is available in <u>the study</u> <u>repository</u>.

## Data Availability

Data in this paper is part of the Human Phenotype Project (HPP) and is accessible to researchers from universities and other research institutions at: https://humanphenotypeproject.org/data-access

https://github.com/mashaashkolnik/causal_framework

## Appendix

### Extended Data

**Extended Data Fig. 1:**
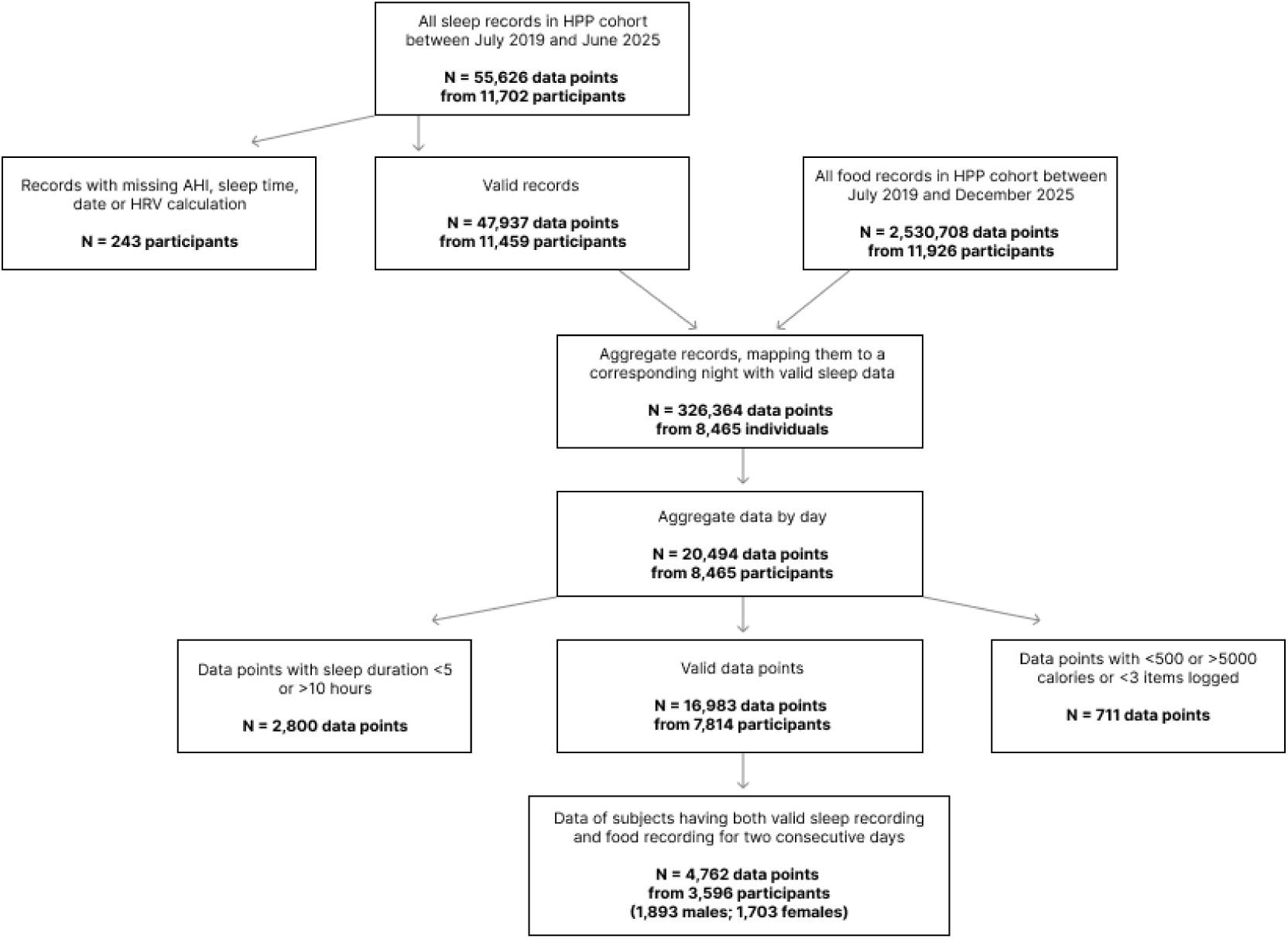
Data Selection and sample sizes. Flow chart showing the different sample sizes of the data available at the time of this work, after the selection process.

**Extended Data Table 1:**
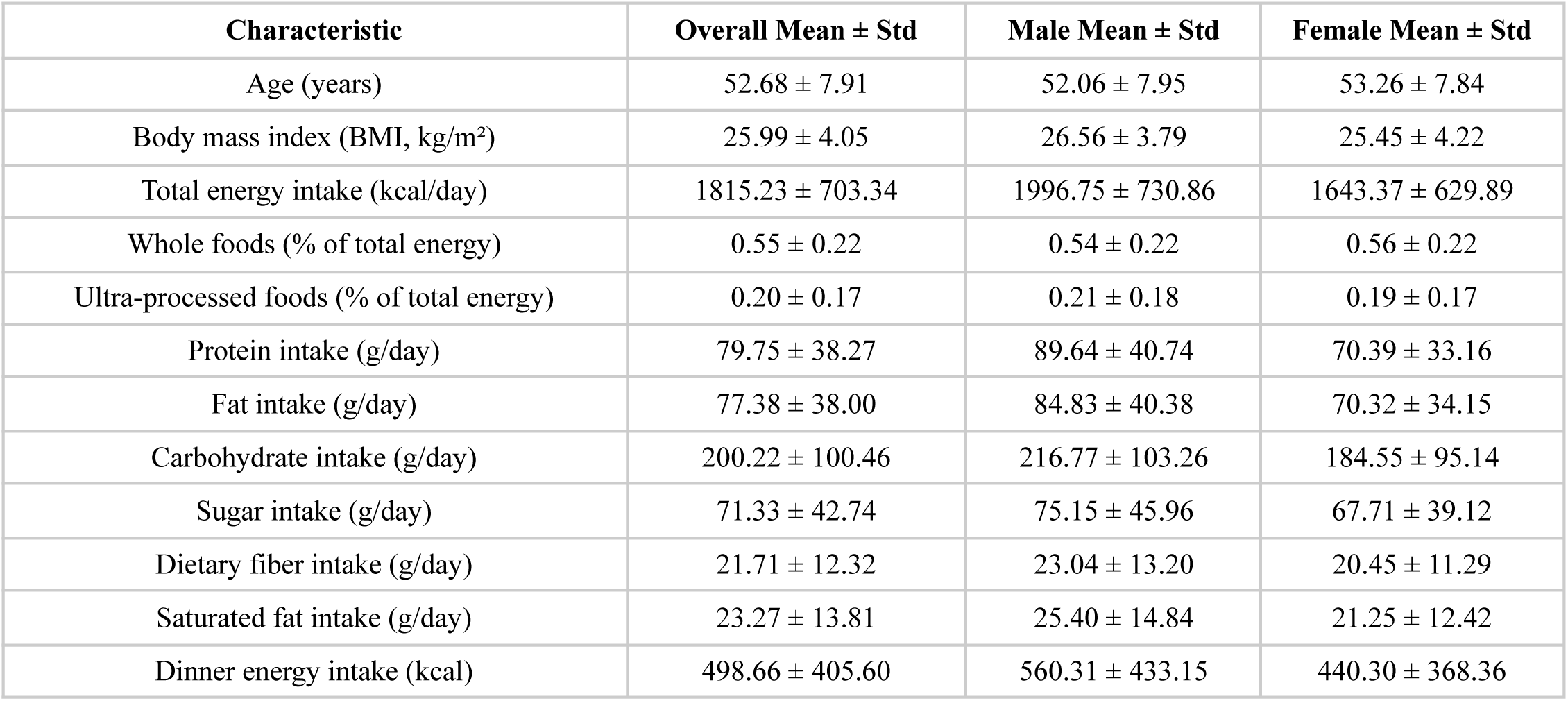
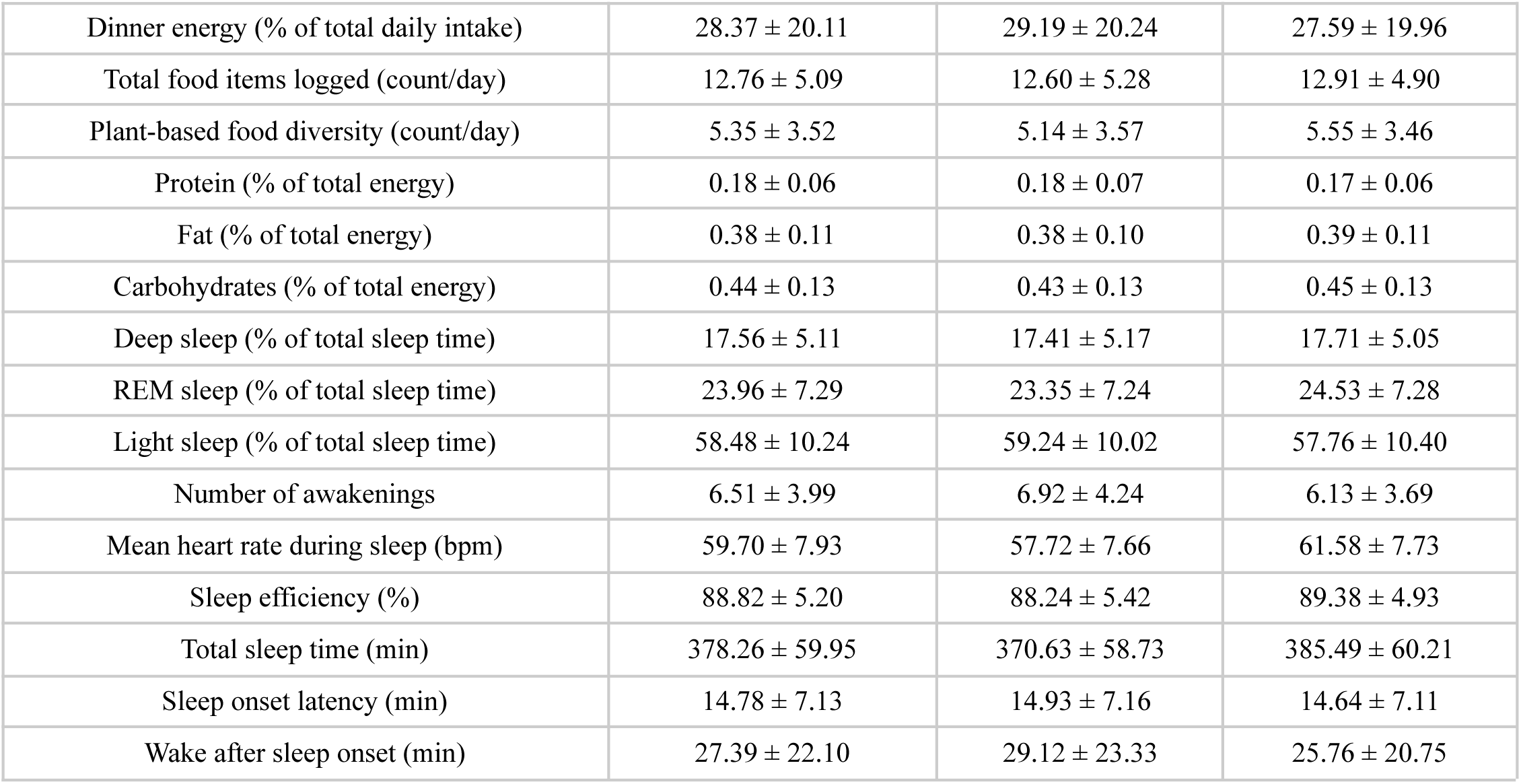
Baseline characteristics of participants included in the analysis. Values are presented as mean ± standard deviation (SD) for the overall cohort and stratified by sex. Dietary variables reflect average daily intake, including total energy, macronutrient composition, food source diversity, and meal timing metrics. Sleep variables were derived from wearable-based nocturnal recordings and summarize sleep architecture, continuity, and autonomic measures.

| Characteristic | Overall Mean ± Std | Male Mean ± Std | Female Mean ± Std |
| --- | --- | --- | --- |
| Age (years) | 52.68 ± 7.91 | 52.06 ± 7.95 | 53.26 ± 7.84 |
| Body mass index (BMI, kg/m<sup>2</sup>) | 25.99 ± 4.05 | 26.56 ± 3.79 | 25.45 ± 4.22 |
| Total energy intake (kcal/day) | 1815.23 ± 703.34 | 1996.75 ± 730.86 | 1643.37 ± 629.89 |
| Whole foods (% of total energy) | 0.55 ± 0.22 | 0.54 ± 0.22 | 0.56 ± 0.22 |
| Ultra-processed foods (% of total energy) | 0.20 ± 0.17 | 0.21 ± 0.18 | 0.19 ± 0.17 |
| Protein intake (g/day) | 79.75 ± 38.27 | 89.64 ± 40.74 | 70.39 ± 33.16 |
| Fat intake (g/day) | 77.38 ± 38.00 | 84.83 ± 40.38 | 70.32 ± 34.15 |
| Carbohydrate intake (g/day) | 200.22 ± 100.46 | 216.77 ± 103.26 | 184.55 ± 95.14 |
| Sugar intake (g/day) | 71.33 ± 42.74 | 75.15 ± 45.96 | 67.71 ± 39.12 |
| Dietary fiber intake (g/day) | 21.71 ± 12.32 | 23.04 ± 13.20 | 20.45 ± 11.29 |
| Saturated fat intake (g/day) | 23.27 ± 13.81 | 25.40 ± 14.84 | 21.25 ± 12.42 |
| Dinner energy intake (kcal) | 498.66 ± 405.60 | 560.31 ± 433.15 | 440.30 ± 368.36 |
| Dinner energy (% of total daily intake) | 28.37 ± 20.11 | 29.19 ± 20.24 | 27.59 ± 19.96 |
| Total food items logged (count/day) | 12.76 ± 5.09 | 12.60 ± 5.28 | 12.91 ± 4.90 |
| Plant-based food diversity (count/day) | 5.35 ± 3.52 | 5.14 ± 3.57 | 5.55 ± 3.46 |
| Protein (% of total energy) | 0.18 ± 0.06 | 0.18 ± 0.07 | 0.17 ± 0.06 |
| Fat (% of total energy) | 0.38 ± 0.11 | 0.38 ± 0.10 | 0.39 ± 0.11 |
| Carbohydrates (% of total energy) | 0.44 ± 0.13 | 0.43 ± 0.13 | 0.45 ± 0.13 |
| Deep sleep (% of total sleep time) | 17.56 ± 5.11 | 17.41 ± 5.17 | 17.71 ± 5.05 |
| REM sleep (% of total sleep time) | 23.96 ± 7.29 | 23.35 ± 7.24 | 24.53 ± 7.28 |
| Light sleep (% of total sleep time) | 58.48 ± 10.24 | 59.24 ± 10.02 | 57.76 ± 10.40 |
| Number of awakenings | 6.51 ± 3.99 | 6.92 ± 4.24 | 6.13 ± 3.69 |
| Mean heart rate during sleep (bpm) | 59.70 ± 7.93 | 57.72 ± 7.66 | 61.58 ± 7.73 |
| Sleep efficiency (%) | 88.82 ± 5.20 | 88.24 ± 5.42 | 89.38 ± 4.93 |
| Total sleep time (min) | 378.26 ± 59.95 | 370.63 ± 58.73 | 385.49 ± 60.21 |
| Sleep onset latency (min) | 14.78 ± 7.13 | 14.93 ± 7.16 | 14.64 ± 7.11 |
| Wake after sleep onset (min) | 27.39 ± 22.10 | 29.12 ± 23.33 | 25.76 ± 20.75 |

**Extended Data Table 2:** Distribution of dietary and meal-timing exposures in treated and control groups. Values are shown as mean ± standard deviation (SD) for treated and control observations, overall and stratified by sex. Exposures include meal timing, dietary composition, food source categories, and dietary diversity metrics. Treatment status reflects binary exposure definitions used in the causal analyses.

| Exposure | Treated<br>Mean ± Std | Control<br>Mean ± Std | Treated<br>Male Mean<br>± Std | Control<br>Male Mean<br>± Std | Treated<br>Female<br>Mean ± Std | Control<br>Female<br>Mean ± Std |
| --- | --- | --- | --- | --- | --- | --- |
| Last-meal-to-bedtime interval (h) | 4.08 ± 1.92 | 2.24 ± 1.43 | 4.03 ± 1.93 | 2.22 ± 1.46 | 4.13 ± 1.90 | 2.27 ± 1.41 |
| Eating window duration (h) | 12.73 ± 2.32 | 9.63 ± 2.58 | 12.82 ± 2.39 | 9.67 ± 2.67 | 12.62 ± 2.23 | 9.60 ± 2.51 |
| Dinner energy (%) | 46.19 ± 18.38 | 29.12 ± 16.14 | 47.08 ± 18.23 | 29.71 ± 16.44 | 45.23 ± 18.49 | 28.59 ± 15.86 |
| Whole foods (% of daily energy) | 0.68 ± 0.18 | 0.41 ± 0.18 | 0.68 ± 0.18 | 0.41 ± 0.18 | 0.69 ± 0.18 | 0.41 ± 0.18 |
| Plant-based foods (% of daily energy) | 0.42 ± 0.19 | 0.18 ± 0.13 | 0.41 ± 0.18 | 0.17 ± 0.13 | 0.43 ± 0.19 | 0.18 ± 0.13 |
| Plant-based food diversity (count) | 7.40 ± 3.35 | 3.06 ± 2.02 | 7.37 ± 3.42 | 2.92 ± 2.02 | 7.44 ± 3.29 | 3.21 ± 2.02 |
| Fruits and vegetables (% of daily energy) | 0.18 ± 0.09 | 0.09 ± 0.07 | 0.17 ± 0.08 | 0.09 ± 0.07 | 0.19 ± 0.09 | 0.10 ± 0.08 |
| Dietary fiber density (g/1,000 kcal) | 15.73 ± 6.01 | 9.10 ± 3.92 | 15.33 ± 5.71 | 8.90 ± 3.88 | 16.05 ± 6.23 | 9.32 ± 3.96 |
| Processed foods (% of daily energy) | 0.29 ± 0.17 | 0.11 ± 0.12 | 0.30 ± 0.17 | 0.11 ± 0.12 | 0.28 ± 0.16 | 0.10 ± 0.12 |
| Animal-based foods (% of daily energy) | 0.35 ± 0.14 | 0.21 ± 0.11 | 0.34 ± 0.14 | 0.21 ± 0.12 | 0.35 ± 0.14 | 0.20 ± 0.11 |
| Dairy (% of daily energy) | 0.13 ± 0.10 | 0.03 ± 0.06 | 0.12 ± 0.10 | 0.03 ± 0.06 | 0.13 ± 0.10 | 0.04 ± 0.07 |
| Meat and poultry (% of daily energy) | 0.17 ± 0.14 | 0.03 ± 0.09 | 0.18 ± 0.14 | 0.04 ± 0.10 | 0.16 ± 0.13 | 0.03 ± 0.08 |
| Saturated Fat (g) | 29.90 ± 13.90 | 16.42 ± 8.77 | 32.73 ± 14.89 | 17.65 ± 9.12 | 27.23 ± 12.33 | 15.26 ± 8.27 |
| Protein (% of daily energy) | 0.21 ± 0.06 | 0.15 ± 0.04 | 0.21 ± 0.06 | 0.15 ± 0.04 | 0.20 ± 0.05 | 0.14 ± 0.04 |
| Fat (% of daily energy) | 0.44 ± 0.09 | 0.32 ± 0.09 | 0.44 ± 0.09 | 0.32 ± 0.08 | 0.44 ± 0.09 | 0.32 ± 0.09 |
| Carbs (% of daily energy) | 0.52 ± 0.10 | 0.37 ± 0.11 | 0.51 ± 0.10 | 0.37 ± 0.11 | 0.52 ± 0.10 | 0.37 ± 0.10 |
| Magnesium (mg) | 485.77 ± 318.76 | 249.09 ± 111.97 | 501.19 ± 277.02 | 268.44 ± 125.10 | 471.17 ± 353.23 | 230.78 ± 94.39 |
| Omega-3 (g) | 2.84 ± 1.85 | 1.37 ± 1.13 | 3.00 ± 1.95 | 1.48 ± 1.19 | 2.68 ± 1.73 | 1.27 ± 1.07 |
| Vitamin D (mcg) | 7.49 ± 9.15 | 2.35 ± 4.41 | 7.60 ± 9.88 | 2.41 ± 4.39 | 7.38 ± 8.41 | 2.29 ± 4.43 |
| Vitamin b6 (mg) | 2.24 ± 1.00 | 1.33 ± 0.66 | 2.51 ± 1.00 | 1.49 ± 0.70 | 1.99 ± 0.94 | 1.19 ± 0.59 |
| Vitamin b12 (ug) | 6.33 ± 9.13 | 2.37 ± 3.15 | 7.25 ± 10.58 | 2.61 ± 3.33 | 5.45 ± 7.41 | 2.14 ± 2.95 |
| Calcium (mg) | 941.96 ± 383.23 | 531.48 ± 305.89 | 965.88 ± 382.13 | 547.05 ± 335.10 | 919.59 ± 383.00 | 516.95 ± 275.10 |
| Zinc (mg) | 11.90 ± 4.72 | 7.32 ± 3.48 | 13.48 ± 4.91 | 8.11 ± 3.83 | 10.43 ± 4.01 | 6.59 ± 2.94 |
| Folic Acid (mcg) | 468.08 ± 240.82 | 261.54 ± 145.26 | 508.08 ± 252.24 | 283.95 ± 156.12 | 430.20 ± 223.01 | 240.34 ± 130.73 |
| Vitamin C (mg) | 154.28 ± 124.43 | 58.17 ± 58.24 | 155.81 ± 112.54 | 57.63 ± 55.74 | 152.84 ± 134.73 | 58.68 ± 60.52 |
| Vitamin E (mg) | 12.46 ± 6.45 | 6.75 ± 4.14 | 13.25 ± 6.62 | 7.14 ± 4.31 | 11.72 ± 6.19 | 6.38 ± 3.95 |

**Extended Data Table. 3:** Definitions of dietary and meal-timing exposures. Percentages represent the proportion of total daily energy intake. Food group classification was based on predefined food-category mappings (see Methods).

| Exposure | Definition |
| --- | --- |
| Last-meal-to-bedtime interval (h) | Time (hours) between the last logged meal ($\geq 15$ kcal) and reported bedtime. |
| Eating window duration (h) | Time (hours) between the first and last logged meals of the day (each $\geq 15$ kcal). |
| Dinner energy (%) | Percentage of total daily energy consumed within the 6 hours preceding bedtime. |
| Whole foods (% of daily energy) | Proportion of total daily energy derived from minimally processed foods, including whole-grain breads and grains, vegetables, fruits, pulses and legumes, nuts and seeds, poultry, fish and seafood, unprocessed red meats, eggs, and oils and fats. |
| Plant-based foods (% of daily energy) | Proportion of total daily energy derived from plant-based sources, including whole-grain breads and grains, vegetables, fruits, pulses, and plant-derived oils and fats. |
| Plant-based food diversity (count) | Number of unique plant-based food items logged per day, including whole grains, vegetables, fruits, pulses, and plant-derived oils. |
| Fruits and vegetables (% of daily energy) | Proportion of total daily energy derived from fruits and vegetables. |
| Dietary fiber density (g/1,000 kcal) | Total dietary fiber intake normalized per 1,000 kcal of total daily energy intake. |
| Processed foods (% of daily energy) | Proportion of total daily energy derived from processed and ultra-processed foods, including fast foods, deep-fried items, processed meats, baked goods, sweetened dairy products, fruit juices and soft drinks, ready-to-eat vegetarian dishes, snacks, low-calorie and diet drinks, sweets, soups, and sauces. |
| Animal-based foods (% of daily energy) | Proportion of total daily energy derived from animal-based foods, including poultry, red and processed meats, eggs, fish and seafood, and dairy products. |
| Dairy (% of daily energy) | Proportion of total daily energy derived from dairy products, including milk, cream, yogurt, soft cheeses, and hard cheeses. |
| Meat and poultry (% of daily energy) | Proportion of total daily energy derived from meat and poultry sources, including poultry, beef, veal, lamb, and other meat products. |

### Supplementary Results

To complement the significant findings presented in the main text, we provide additional analyses of nutritional exposures that did not yield statistically significant associations with next-night objective sleep physiology after bootstrap-based uncertainty estimation and inverse-probability weighting. These results help illustrate the specificity of our causal effects framework and highlight dietary domains where day-to-day nutritional variation did not produce measurable short-term changes in sleep architecture. For transparency, all outcomes are shown with 95% confidence intervals.

**Supplementary Results Figure S1.**
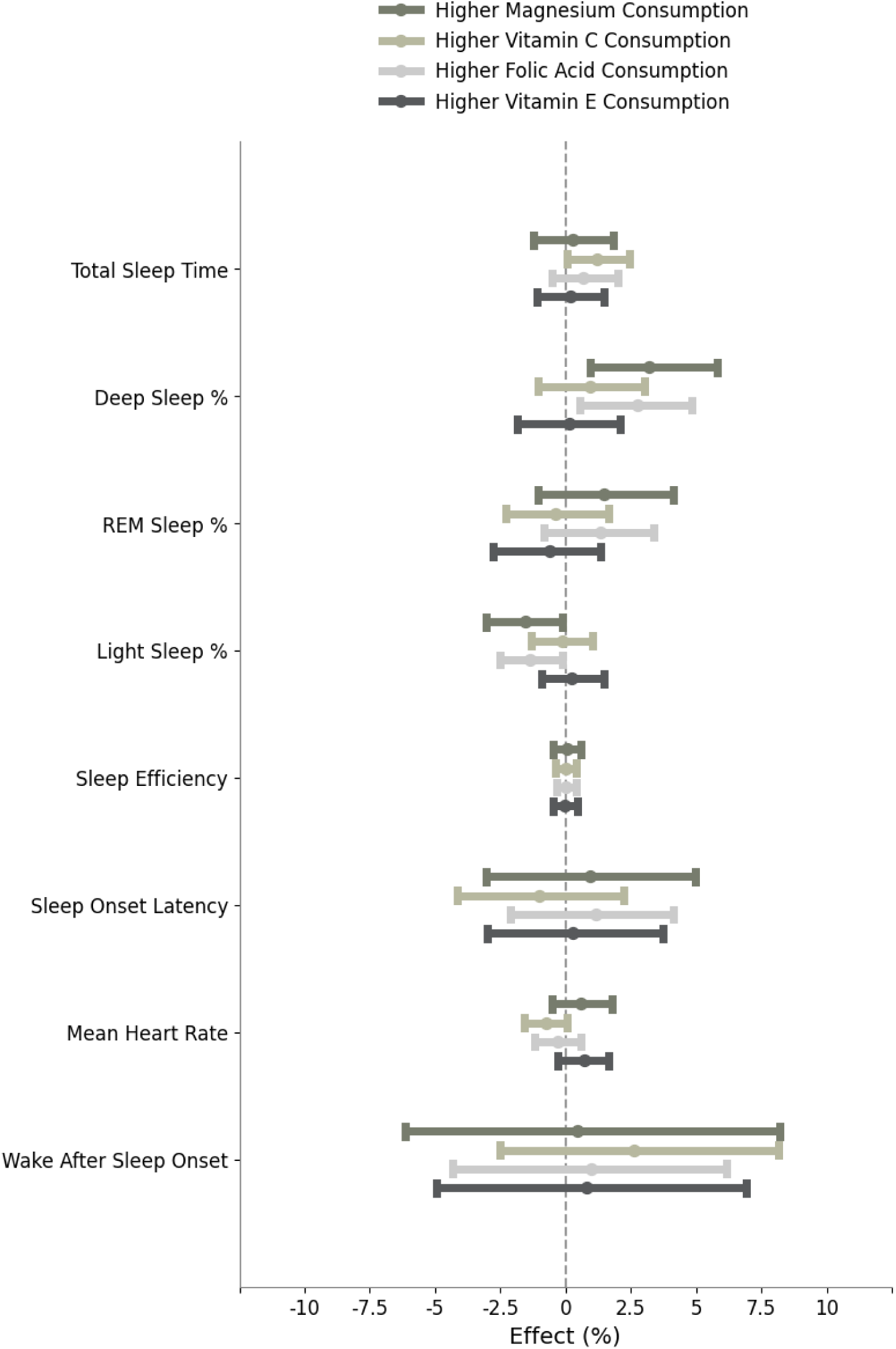
Plant-Associated Vitamins Show Consistent Trends Toward Improved Sleep Architecture, Despite Limited Statistical Significance. This figure presents inverse-probability–weighted average treatment effects (ATEs) comparing higher versus lower daily intake of plant-correlated micronutrients (magnesium, vitamin C, folate, and vitamin E) on objective sleep outcomes. Error bars represent 95% bootstrap confidence intervals. Across several sleep domains, including deep sleep percentage, REM sleep percentage, and total sleep time, point estimates were directionally consistent and suggestive of beneficial associations with higher intake of these vitamins. Notably, for several outcomes the confidence intervals did not cross the null, indicating relatively stable effect estimates across bootstrap resamples. However, corresponding p-values remained above the nominal significance threshold (p > 0.05) after the correction.

**Supplementary Results Figure S2.**
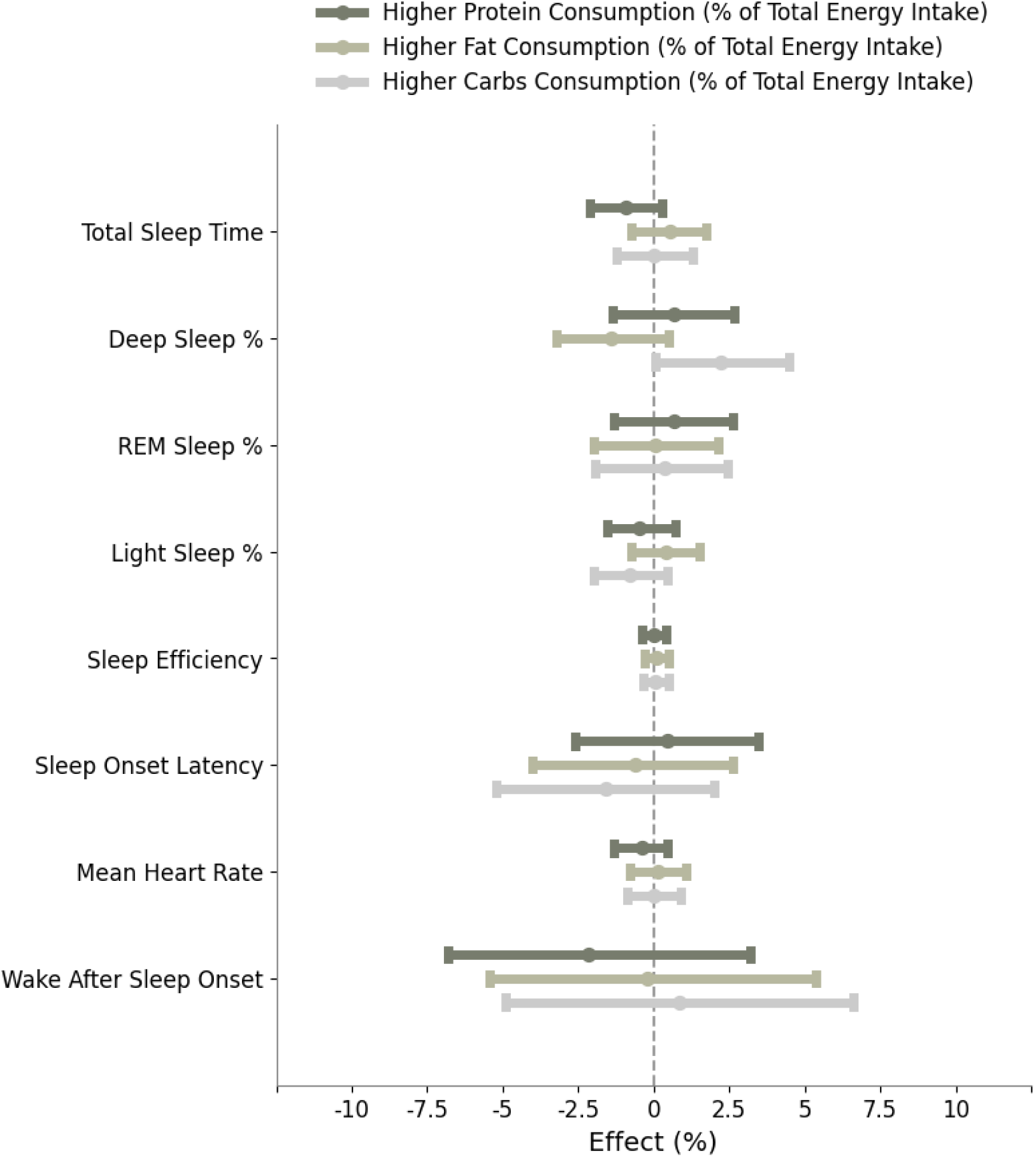
Macronutrient Energy Distribution Shows No Robust Short-Term Effects on Objective Sleep Physiology. This figure presents inverse-probability–weighted average treatment effects (ATEs) comparing higher versus lower proportional intake of protein, fat, and carbohydrates (expressed as percentage of total daily energy intake) on objective sleep outcomes. Error bars denote 95% bootstrap confidence intervals. Across all sleep metrics, including sleep duration, sleep stage composition, sleep efficiency, nocturnal heart rate, sleep onset latency, and wake after sleep onset, none of the macronutrient contrasts yielded statistically significant associations (all p > 0.05) after correction.

**Supplementary Results Figure S3.**
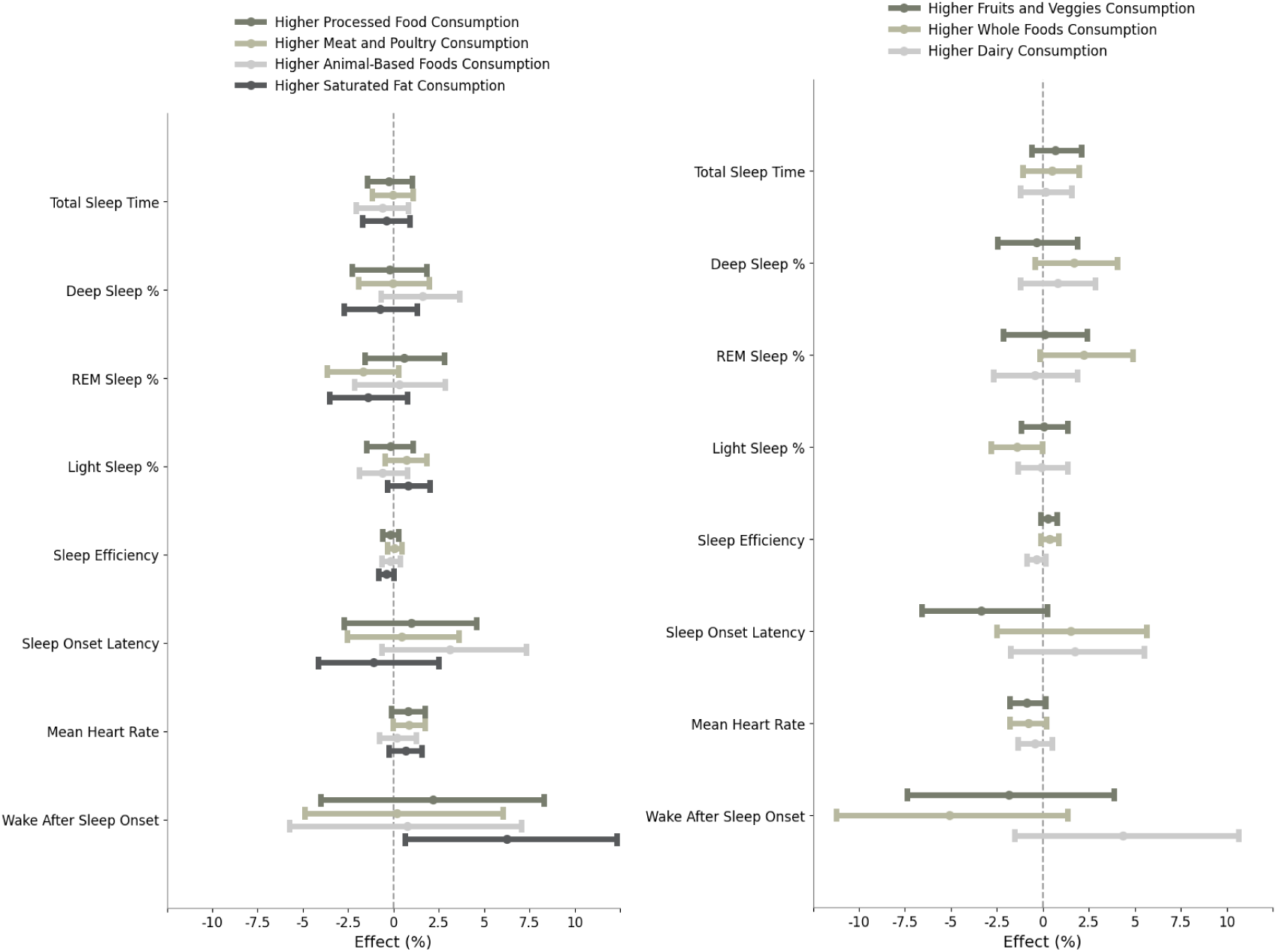
Food-Source-Specific Energy Patterns Show Divergent Associations with Objective Sleep Physiology. The figure shows inverse-probability–weighted average treatment effects (ATEs) comparing higher versus lower intake of food-source–specific dietary patterns, expressed as energy ratios or percentage of total energy intake. The left panel summarizes processed food, meat and poultry, animal-based whole foods, and saturated fat intake, while the right panel summarizes plant-forward patterns, including fruits and vegetables, whole plant foods, whole foods overall, and dairy. Error bars denote 95% bootstrap confidence intervals. Across outcomes, several exposures exhibited confidence intervals that did not cross the null, indicating statistically robust associations at the confidence-interval level. Importantly, the direction of effects varied systematically by food source. Higher intake of plant-based and whole-food patterns tended to be associated with favorable sleep characteristics, including higher proportions of deep and REM sleep and reduced wake after sleep onset. In contrast, higher intake of processed foods, saturated fat, and certain animal-based energy ratios showed associations in the opposite direction, including longer wake after sleep onset and less favorable sleep-stage distributions.

### Extreme intake contrasts

Prior work examining lifestyle-sleep relationships has shown that increasing the magnitude of behavioral contrast can uncover stronger physiological effects that are attenuated under population-average comparisons. For example, a recent study demonstrated dose-dependent disruptions in sleep and nocturnal autonomic function by contrasting progressively higher levels of evening exercise strain rather than relying on binary exposure definitions ^68^. Motivated by this framework, we performed complementary contrast-amplification analyses for all dietary and behavioral exposures that were significant in the primary median-based comparisons.

Specifically, for each shortlisted exposure, including meal timing (hours to sleep, eating window), caloric timing (nighttime caloric intake), dietary composition (plant-based and animal-based food ratios, whole and processed food categories), dietary diversity, and fiber density, we contrasted individuals in the top and bottom 30% of the exposure distribution using 1:1 propensity-score matching. Matching was performed on baseline dietary intake, demographic variables, and anthropometric measures, using a caliper of 0.15 standard deviations of the logit of the propensity score to ensure covariate balance while increasing separation between exposure groups. Across exposures, this design yielded approximately 200-300 matched pairs per comparison. Statistical significance was assessed using covariate-adjusted mean differences estimated via CUPED ^69^, followed by two-sample t-tests on matched pairs, with false discovery rate correction applied across outcomes.

Compared with the primary analysis, extreme-quantile matching resulted in reduced sample sizes and less stringent covariate balance, with mean post-matching absolute standardized mean differences. Given these limitations, we present the extreme-quantile results in the Appendix as supportive analyses rather than primary evidence.

Despite reduced power, effect directions were consistent with the main results across exposures. Notably, fiber density continued to show statistically significant differences in sleep-stage composition and nocturnal heart rate under extreme contrasts, with larger effect magnitudes than observed in the median-based analysis (Appendix Fig. S4). Other dietary features, including whole-food and processed-food categories, did not reach statistical significance under extreme-quantile matching; however, their estimated effects remained directionally consistent with the primary analysis, suggesting that limited power rather than effect reversal likely explains the absence of significance.

**Supplementary Results Figure S4.**
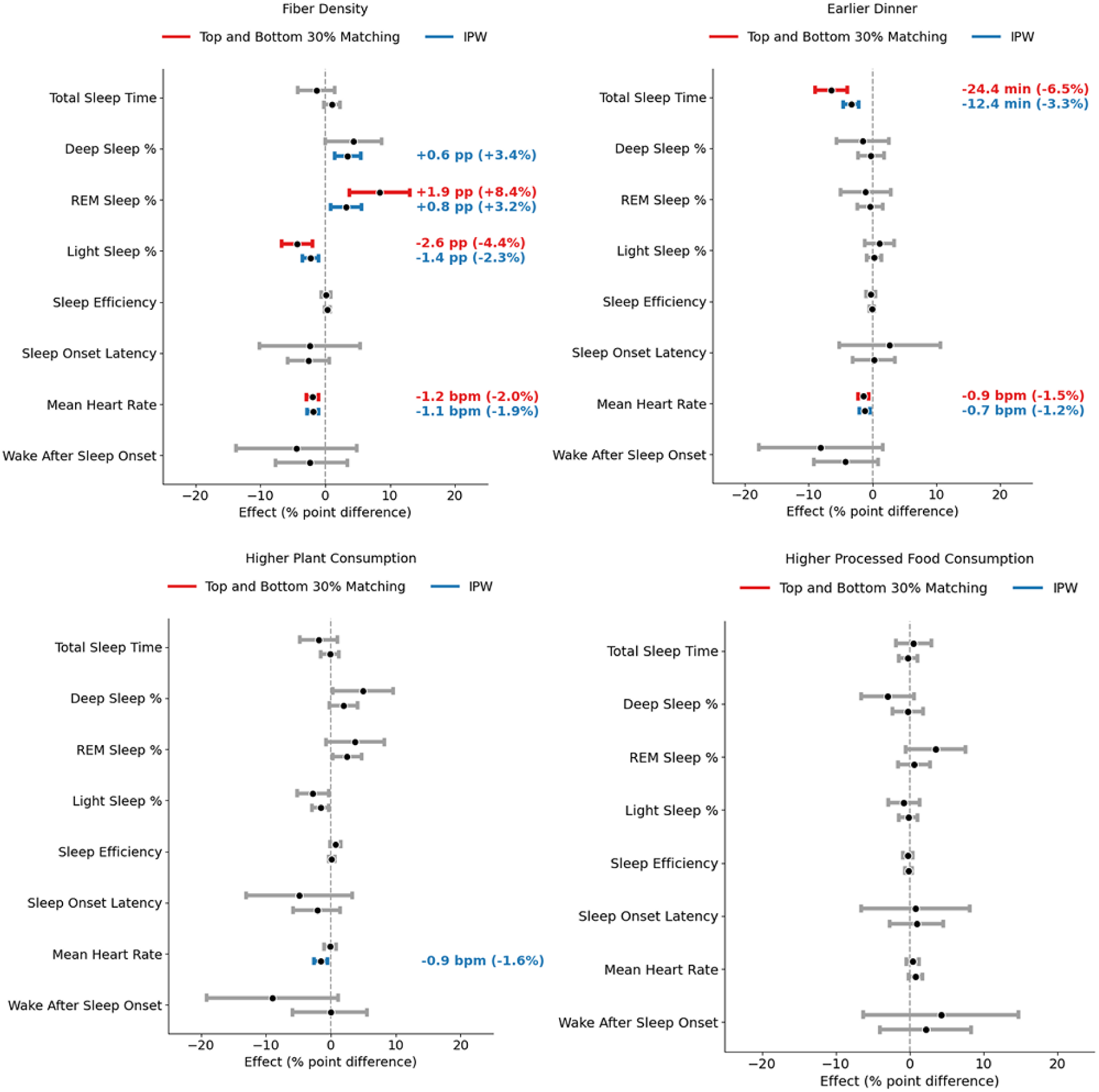
Extreme-quantile matching reveals amplified but less precisely estimated diet-sleep associations. This figure presents average treatment effects estimated after 1:1 propensity-score matching between individuals in the top and bottom quartiles (25%) of selected dietary and behavioral exposures, including fiber density, dinner timing, whole plant food intake, and processed food intake. Points indicate estimated effects on next-night sleep outcomes expressed as percent differences relative to matched controls, with horizontal bars denoting 95% confidence intervals.

